# A cell-level discriminative neural network model for diagnosis of blood cancers

**DOI:** 10.1101/2023.02.07.23285606

**Authors:** Edgar E. Robles, Ye Jin, Padhraic Smyth, Richard H. Scheuermann, Jack D. Bui, Huan-You Wang, Jean Oak, Yu Qian

## Abstract

**Motivation:** Precise identification of cancer cells in patient samples is essential for accurate diagnosis and clinical monitoring but has been a significant challenge in machine learning approaches for cancer precision medicine. In most scenarios, training data are only available with disease annotation at the subject or sample level. Traditional approaches separate the classification process into multiple steps that are optimized independently. Recent methods either focus on predicting sample-level diagnosis without identifying individual pathologic cells or are less effective for identifying heterogeneous cancer cell phenotypes.

**Results:** We developed a generalized end-to-end differentiable model, the Cell Scoring Neural Network (CSNN), which takes the available sample-level training data and predicts both the diagnosis of the testing samples and the identity of the diagnostic cells in the sample, simultaneously. The cell-level density differences between samples are linked to the sample diagnosis, which allows the probabilities of individual cells being diagnostic to be calculated using backpropagation. We applied CSNN to two independent clinical flow cytometry datasets for leukemia diagnosis. In both qualitative and quantitative assessments, CSNN outperformed preexisting neural network modeling approaches for both cancer diagnosis and cell-level classification. Post hoc decision trees and 2D dot plots were generated for interpretation of the identified cancer cells, showing that the identified cell phenotypes match the cancer endotypes observed clinically in patient cohorts. Independent data clustering analysis confirmed the identified cancer cell populations.

**Availability:** The source code of CSNN and datasets used in the experiments are publicly available on GitHub and FlowRepository.

**Contact:** Edgar E. Robles: and Yu Qian

**Supplementary information:** Supplementary data are available on GitHub and at *Bioinformatics* online.

## 1 Introduction

Challenges in the diagnosis and prognosis of blood cancers partially lie in the phenotypic heterogeneity of cancer cells. Even within a leukemia subtype, leukemic cells may be derived from slightly different stages of the normal cell developmental trajectory and therefore express different marker proteins, resulting in phenotypic heterogeneity within and between patient samples. To characterize the cellular phenotypic heterogeneity, single-cell assays are essential. In clinical laboratories, complete blood cell count (CBC), cytogenetics for identifying chromosomal numerical and structural abnormalities, microscopy of bone marrow biopsy, cytology of cerebrospinal fluid, and flow cytometry (FCM) of peripheral blood and bone marrow aspirates are commonly used for leukemia and lymphoma diagnosis. Among these assays, FCM is the most mature single cell analysis technology, supporting identification and quantification of cell surface and intracellular proteins on individual cells. Compared with other single cell assays, FCM is rapid, cheap, and sensitive for detecting and monitoring phenotypic differences in cancer cells. Besides profiles of cellular marker expressions, proportions of the cancer cells within a specimen (i.e., cancer burden, an important measure for optimizing treatments and prognosis) can also be quantified from the analysis of FCM data. As a result, FCM immunophenotyping is routinely used in diagnosis and prognosis of blood and lymphoid cancers [6, 55, 46, 56, 60, 22, 10, 23, 54, 7]. It is also widely used to identify abnormal cell populations in association with non-neoplastic diseases including asthma, allergy, and autoimmunity [52, 19, 8, 4, 61, 25, 59].

Due to significant advances in cytometry instrumentation and reagent technologies since the 2000s [53, 39, 20, 41], applications of cutting-edge machine learning (ML) approaches began to emerge in the recent decade for addressing the increasing volume and complexity in this high-content cytometry data [18, 33, 47]. Automated gating analysis (auto-gating), which identifies cell populations using unsupervised clustering methods or recapitulates the manual identification gating process using supervised learning, in either the original or transformed feature space [29, 43, 36, 9, 63, 40, 44, 38, 34, 12, 48, 2, 26, 28, 57, 31, 1, 45, 51, 27], represents the largest category of these methods. To use these auto-gating methods for diagnosis, a separate disease classification step is required. The accuracy of the classification relies on the auto-gating step to identify cell populations in a complete and accurate way, which can be challenging for blood cancer applications. A second category of methods focuses on identifying cell-based biomarkers from the FCM data by extracting statistical features from cell-level expression patterns and comparing them between samples [30, 15, 5, 13, 58, 62]. The identified biomarkers are selected to be statistically different between cohorts but may not be biologically meaningful cell populations with distinct phenotypes. These methods focus on identifying cohort-level differences and are not designed for or dependent on the accurate identification of (cancer) cells in each individual sample. A third category of methods makes use of representation learning models, such as neural networks, to bypass the feature extraction step. Some of these methods are shown to be effective in predicting the cancer diagnosis [37, 24] but the predicted diagnosis is difficult to interpret and validate without identifying the cancer cells themselves. Another group of methods in this category has been applied to non-neoplastic diseases for predicting the sample diagnosis while identifying the diagnostic cell populations [16, 3, 17]. However, the identified cell populations were not validated due to the lack of cell-level labels and it remains unclear whether these methods can effectively identify cancer cell populations with phenotypic heterogeneity.

Here we define the problem to be solved as follows. Given a set of preexisting clinical FCM samples with diagnostic labels, with cancer burden being optionally available (as identified by expert manual or automated gating analysis), can we predict the diagnosis label of a new FCM sample from the same reagent panel and identify the diagnostic cells simultaneously, while retaining interpretability of the identified cancer cells. Addressing this problem requires the simultaneous optimization of cell population identification (biomarkers) and sample-level classification (diagnosis). In previous work [21], we showed that the simultaneous optimization can be achieved for diagnosis of chronic lymphocytic leukemia (CLL), by adapting gradient descent optimization for identifying a global optimal gate to maximize classification accuracy. In this paper, we hypothesize that a density-based discriminative point set model using backpropagation can address the simultaneous optimization, without requiring initial gating.

Specifically, we developed an end-to-end differentiable representation learning approach - Cell Scoring Neural Network (CSNN) - that learns the density distribution of the cellular expressions on all markers and makes diagnostic predictions based on aggregating cell-level scores into sample-level predictions. In parallel, the sample-level information, including the diagnostic labels and the density patterns, is backpropagated to the cell level for identifying the diagnostic cell population(s). Based on the possible availability of cancer burden information, we developed two versions of CSNN: CSNN-Class (classification), which requires only diagnostic labels in the training data and CSNN-Reg (regression), which makes use of the additional sample-level cancer burden information to improve the prediction at the single cell level. We applied both CSNN modeling methods on two independent datasets for diagnosis of CLL (provided by University of California, San Diego) and B-ALL (B-cell lymphoblastic leukemia, provided by Stanford University). We compared the performance of the resulting models with two relevant representative deep learning modeling approaches, cellCNN [3] and DeepCellCNN [17], assessing their clinical utilities regarding: a) accuracy of diagnostic prediction, b) interpretability of the identified leukemic cell populations and their phenotypic heterogeneity, and c) accuracy of the identified cancer burden. To confirm that the CSNN-identified diagnostic cell populations are those that differ between the cancer and non-cancer samples, we designed and performed independent data clustering analysis to identify the clusters of cells that can only be found from the cancer samples and compared them with the CSNN-identified cancer cells. To interpret the CSNN-identified phenotypic heterogeneity of the cancer cells, we constructed post hoc decision trees based on the predicted cell-level labels, enumerated all leukemic cell phenotypes along the tree paths, and compared them with the known cancer endotypes observed clinically in patient cohorts. The leukemic cell populations identified in individual samples are then visualized in traditional 2D dot plots for straightforward hematopathology review.

## 2 Methods

### 2.1 Notation

We consider *N* individuals (e.g., patients) where each individual *i*, 1≤ *i*≤ *N*, is represented by an FCM sample *X*_*i*_. Each sample *X*_*i*_ consists of a set of multi-dimensional vector measurements, where each vector *x*_*i,j*_ in the sample corresponds to a single cell, i.e., 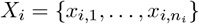, where *j* is an index of cells in sample *X*_*i*_, *n*_*i*_ is the number of cells in sample *X*_*i*_, and each dimension corresponds to the expression of an FCM marker.

We assume a target *y*_*i*_ is available for each subject *i*, provided (for example) by human experts based on manual evaluation of the FCM sample *X*_*i*_. In this paper, we will consider two different types of targets *y*_*i*_. The first type of target is a real-valued target *y*_*i*_, taking values between 0 and 1, indicating the disease burden for sample *i*, i.e., the proportion of cells that are estimated to be pathogenic for that sample, with *y*_*i*_ = 0 for healthy samples and *y*_*i*_ *>* 0 for samples diagnosed with the disease condition. The second type is a binary target *y*_*i*_, taking the value 0 or 1, indicating a disease diagnosis for sample *i*, and where *y*_*i*_ = 1 indicates disease presence for sample *i*.

At the cell level, let *z*_*i,j*_ be a cell-level binary variable where *z*_*i,j*_ = 1 indicates that the cell is pathogenic and *z*_*i,j*_ = 0 indicates that a cell is non-pathogenic (present in healthy conditions). We will assume that cell-level labels are not available in the training data, i.e., that the *z*_*i,j*_ value for cell *j* for patient *i* is unknown.

An important aspect of our overall approach is to be able to predict, for patient *i*, both the cell-level binary variables *z*_*i,j*_ and the sample-level disease diagnosis *y*_*i*_. More specifically, we estimate both:

1. **cell-level scores** *s*_*i,j*_, in the form of conditional probabilities *s*_*i,j*_ = *P* (*z*_*i,j*_ = 1| *x*_*i,j*_), i.e., the probability that a particular cell is pathogenic, given marker measurements *x*_*i,j*_; and
2. **sample-level** real-valued burdens *y*_*i*_ where these sample-level estimated burden is a functions of the cell-level scores *s*_*i,j*_.

### 2.2 A Cell-Scoring Neural Network for Disease Prediction

Our goal is to construct a predictive model that takes a set of cell-level vectors for a sample, 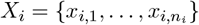, and produces a sample-level prediction of *y*_*i*_ (either a real-valued burden or a probability of a binary label). A challenge in this context is that predictive modeling techniques in statistics and machine learning typically assume a fixed-dimensional vector representation as input to a model, rather than sets of vectors *X*_*i*_ of varying sizes across *i*.

To handle this issue, we use the following two-step approach. In the first step we map each cell-level vector *x*_*i,j*_ to a scalar-valued cell-level conditional probability score *s*_*i,j*_ = *P* (*z*_*i,j*_ = 1 |*x*_*i,j*_) where the mapping *s*_*i,j*_ = *s*(*x*_*i,j*_; *ϕ*) has learnable parameters *ϕ*, parametrized via a feedforward neural network, which we refer to as a Cell Scoring Neural Network (CSNN). Using this cell-level mapping, each sample 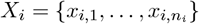 can then be represented by a set of cell-level scores 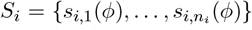, where each score indicates how likely it is that a particular cell *i, j* is pathogenic. Note that the cell-level scores *s*_*i,j*_(*ϕ*) depend implicitly on the cell-level data vectors *x*_*i,j*_; we suppress this dependence on *x*_*i,j*_ in the notation for simplicity.

In the second step, to predict real-valued burden targets *y*_*i*_, we aggregate the cell-level scores by averaging, i.e., 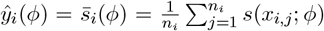, representing an estimate of disease burden for sample *X*_*i*_. To predict a binary target we define *P* (*y*_*i*_ = 1|*X*_*i*_) to be a logistic function, i.e.,

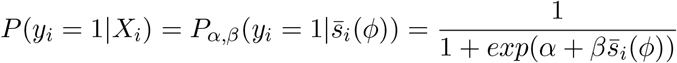

where *α* and *β* are learnable parameters of the logistic function.

Note that the two types of predictions have different interpretations. The burden prediction *y*_*i*_ can in practice take values quite close to 0 for patients who have a disease diagnosis (e.g., a patient could have as few as 0.01% pathogenic cells and still have the disease). On the other hand, the conditional probability estimate, *P* (*y*_*i*_ = 1 |*X*_*i*_), can be interpreted as having a threshold at 0.5, i.e., if *P* (*y*_*i*_ = 1 |*X*_*i*_) *>* 0.5 then individual *i* is more likely to have the disease than not (and vice-versa). The logistic parameters allow for accommodation of this difference between predicting burden level and predicting likelihood of disease presence.

A key feature of our approach is that we use information at the sample-level (the *y*_*i*_’s) to learn the mappings for the cell-level scores (the *s*_*i,j*_’s). In particular, for real-valued burden targets we pursue a regression approach and minimize a weighted mean-square error loss function:

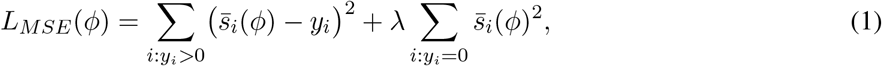

where *λ >* 1 is a hyperparameter that upweights the second term to encourage the model to push the predictions (burden estimates) for healthy individuals to be close to 0.

For binary targets *y*_*i*_∈ {0, 1}, we estimate the parameters by minimizing the standard binary cross-entropy objective function used in classification modeling [14]:

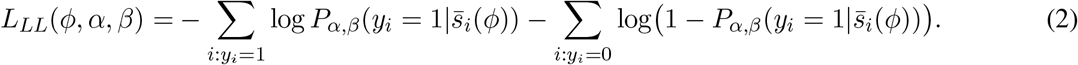

We learn *ϕ* for *L*_*MSE*_ (and simultaneously, *ϕ, α* and *β* for *L*_*LL*_) by using standard gradient descent optimization methods. In what follows, we refer to the first approach above (with real-valued burdens and squared error functions) as CSNN-Reg (for cell scoring neural network regression), and the second approach (with binary labels and log-loss functions) as CSNN-Class (for cell scoring neural network classification).

To represent the cell-level mappings, *x*_*i,j*_ →*s*_*i,j*_(*ϕ*), for both CSNN-Reg and CSNN-Class, we use a flexible function approximator in the form of a multi-layer feedforward neural network. In particular we use a ReLU activation function in the intermediate hidden layers and a sigmoid (softmax) function, *g*(*z*) = 1*/*(1 + exp(− *z*)) as the activation function at the output layer so that the model’s output per cell is constrained to lie between 0 and 1. Additional details on network architectures and hyperparameter settings for optimization are provided in *Appendix 1*.

CellCNN [3] and DeepCellCNN [17] are two existing methods that are comparable to a CSNN since they use neural networks that produce multiple scores per cell, which are then aggregated for sample-level classification. In these methods, the scoring neural network is replaced by a neural network that maps every cell to a vector in a latent space, rather than a single score. These vectors are then averaged together, into a final feature vector which is an abstract representation of the sample. This feature vector serves as a summary of the samples, but is difficult to interpret by humans.

In contrast, a distinct advantage of the interpretable cell-level scores produced by the CSNN models is that for any cell, for a particular sample, we can explore and interpret where pathogenic cells are located in marker space, e.g., by visualizing and highlighting what regions in marker space have scores *s*_*i,j*_ = *s*(*x*_*i,j*_; *ϕ*) above a particular threshold. We discuss how this is related to the concept of manual “gating” in *Appendix 2* and provide illustrations of how this can support biologically-meaningful discovery with real FCM datasets later in the paper.

### 2.3 Initializing a CSNN Models using Density Estimates

The quality of the learned CSNN models can be improved by using information related to the densities in marker space to initialize the models. We begin by generating initial estimates of cell-level scores 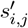 for each cell *j* in each sample *i* in the training data. Consider first the case of real-valued *y*_*i*_ targets (i.e., sample burdens). For samples with *y*_*i*_ = 0, all cell-level scores are zero by definition, i.e., 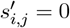, assuming that all cells in healthy samples are non-pathogenic. For samples with a disease diagnosis (*y*_*i*_ *>* 0) Bayes’ rule is used:

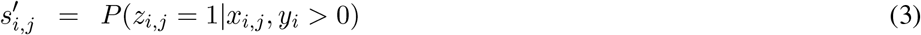

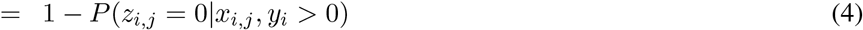

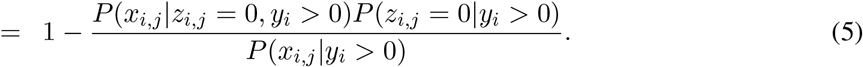

We can estimate each of the three terms on the right hand side from the training data as follows. The term in the denominator, *P* (*x*_*i,j*_| *y*_*i*_ *>* 0), is the marginal probability density function (PDF) of marker measurements for sample *i*, given that sample *i* has a disease diagnosis: this PDF can be estimated straightforwardly using kernel density estimation (KDE). The first term in the numerator, *P* (*x*_*i,j*_ |*z*_*i,j*_ = 0, *y*_*i*_ *>* 0) is the probability density for non-pathogenic cells in a sample with a disease diagnosis. We can approximate this by assuming that *P* (*x*_*i,j*_| *z*_*i,j*_ = 0, *y*_*i*_ *>* 0) = *P* (*x*_*i,j*_| *z*_*i,j*_ = 0) = *P* (*x*_*i,j*_ |*z*_*i,j*_ = 0, *y*_*i*_ = 0), i.e., that the PDF of non-pathogenic cells in marker space is the same in both positive and negative samples. We further assume that the density *P* (*x*_*i,j*_ |*z*_*i,j*_ = 0, *y*_*i*_ = 0) does not vary from sample to sample, allowing us to pool all *x*_*i,j*_ measurements from all the negative samples (which have *y*_*i*_ = 0 and *z*_*i,j*_ = 0 by definition) and again use KDE to estimate this density. The second term in the numerator, *P* (*z*_*i,j*_ = 0| *y*_*i*_ *>* 0), is equal to 1 −*y*_*i*_, under the assumption that *y*_*i*_ (the sample burden) corresponds to the fraction of cells in sample *i* that are pathogenic. For the situation where the training data only contains binary labels *y*_*i,j*_∈ {0, 1}, i.e., the CSNN-Class model, we again estimate scores 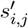 via Bayes rule, but using additional approximations for each of the three required terms in the absence of known burdens *y*_*i*_.

Finally, given scores 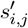 from the density-based approach above (for all cells for all samples in the training dataset), a feedforward neural network, parametrized by weights *ϕ*^*′*^, is trained to create an initial cell-level model that can approximate the density-based scores via the neural network. The trained weights *ϕ*^*′*^ of this neural network are then used to initialize the weights in training of an end-to-end CSNN network (CSNN-Reg or CSNN-Class, using *L*_*MSE*_ and *L*_*LL*_ respectively as described earlier). We found in practice that this density-based initialization significantly improves the quality of the final sample-level disease predictions. Full details on KDE methods, approximations for binary *y*_*i*_ labels, and training of the initial network, are provided in the Supplement.

Note that one could in principle use the density-based approach alone to build a sample-level prediction model, by using the density-based models from the training to generate scores 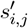 per cell for a new sample. A prediction for *y*_*i*_ could then be based, for example, on thresholding the sum of cell-level scores 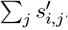. However, a purely density-based approach may be sensitive to modeling assumptions, whereas the sample-level discriminative training of the CSNN can allow the model to further tune the initial parameters *ϕ*^*′*^ to produce scores *s*_*i,j*_ that are directly optimized for robust prediction of burden or likelihood of disease.

## 3 Experiment Results

### 3.1 Datasets

Two independent FCM datasets were used in evaluating the performance of the CSNN modeling methods. The first dataset (DS1) was provided by the University of California, San Diego (UCSD) Center for Advanced Laboratory Medicine (CALM) diagnostic lab that was collected and analyzed for the identification of Chronic Lymphocytic Leukemia (CLL) cases using their standard diagnostic protocol. DS1 includes FCM data from 288 subjects - 186 diagnosed as CLL and 102 judged to be non-CLL by the hematopathologist. For each subject, two reagent panels, PB1 and PB2 were used in the clinical FCM assay on peripheral blood (PB) samples. Each panel contained antibodies for the detection of 10 markers (fluorescence parameters): PB1: CD3, CD5, CD10, CD19, CD22, CD38, CD43, CD45, CD79b and CD81; PB2: Anti-Ig-lambda, Anti-Ig-kappa, CD5, CD7, CD19, CD20, CD23, CD38, CD49d and FMC-7. Datasets from each panel also included 6 scatter parameters: forward scatter (FSC)-area(A)/height(H)/width(W) and side scatter (SSC)-A/H/W.

The second FCM dataset (DS2) was provided by the diagnostic lab at Stanford University (Stanford) for the identification of leukemic cells (blasts) of B-cell Acute Lymphoblastic Leukemia (B-ALL) using their standard diagnostic protocol. The samples in DS2 are bone marrows, which consist of both diagnostic samples and samples collected from patients who have received CD19-targeted CAR (chimeric antigen receptors) T-cell therapy. DS2 includes FCM data from 178 subjects - 50 diagnosed as B-ALL and 128 judged to be non-B-ALL. The FCS files of DS2 are from one reagent panel detecting 9 markers: CD66b, CD22, CD19, CD24, CD10, CD34, CD38, CD20, CD45, and 2 scatter parameters (FSC/SSC).

Research on both datasets was approved by Institutional Review Boards of the respective institutions (UCSD and Stanford). Both datasets have been fully de-identified before being transferred and analyzed using the proposed neural networks. Diagnostic labels of DS1 samples were provided by UCSD. Cancer burden of each DS1 sample was obtained by applying the DAFi automated gating analysis [26] on the de-identified FCS files, following the gating strategy used in the diagnostic lab (*Supplementary Figure 1*). Specifically, DAFi examined the property of all markers, focusing on identifying the CD5^+^CD19^+^CD10^−^CD79b^dim^ CLL cells in PB1 using data clustering, according to our previous study [49]. For DS2, both the diagnosis and the cancer burden were provided by Stanford from expert manual gating analysis.

### 3.2 Performance assessment

#### 3.2.1 Training and testing sets

To assess the performance of the ML modeling approach developed, each dataset was divided into separate subsets for training, validation, and testing. The CLL dataset (DS1) is divided into a training set of size 102, a validation set of size 81, and a testing set of size 105. For the B-ALL dataset (DS2), 118 out of the 178 samples were selected at random and reserved as testing samples; the models were trained with the remaining 60 samples. Using the 60 training samples, hyperparameter values were optimized by running 5-fold cross validation runs.

#### 3.2.2 Quantitative assessment of classification accuracy

The performance of CSNN-Class and CSNN-Reg methods on the CLL and B-ALL datasets were compared with with two machine learning methods recently reported in the literature, CellCNN and DeepCellCNN. For each method hyperparameter optimization was performed by learning parameters on the training subsets and selecting the best model parameter setting based on the area under the receiving operating characteristic curve (AUROC) obtained with the validation subsets. We then retrained the best performing model on a sample containing all the samples in the training set and validation set and determined the final performance on unseen test subsets.

Hyperparameters evaluated for all models included the learning rate and an architecture search, with specific searches for *w* (CSNN-Class), *λ* (CSNN-Reg) and the dropout rate (CellCNN). The specific grid values tested for these can be found in the *Appendix 1*. Using the best hyperparameters for each model, we then trained 20 more models of each type with different random initializations to measure their variance with respect to initialization. In order to filter out all the models that initialized incorrectly we ran each training loop with 5 restarts and evaluated them with the testing set, as described in Fig 2. We then picked the best run out of those 20 to report the ROC graphs in Fig 2. Overall, the proposed CSNN-Reg and CSNN-Class produce higher AUROC scores than either CellCNN or DeepCellCNN. CSNN-Reg is superior to all methods on both datasets, indicating that the sample-level burden information (as used by this method) carries additional value beyond sample-level binary labels (as used by the other three methods). In addition, both of the CSNN models are more robust than the other methods in that they have lower variance (than the other methods) across weight initializations during model training.

**Fig. 1:**
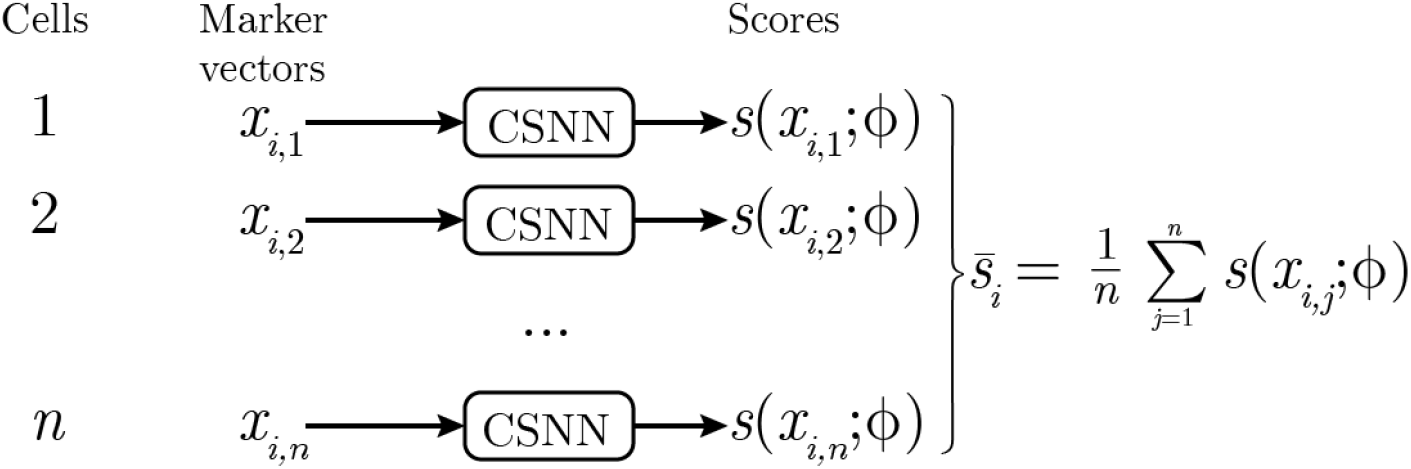
Sample-level predictions from cell-level scores. Each marker vector is evaluated by the CSNN and given a score *s*(*x*_*i,j*_; *ϕ*) that corresponds to *P* (*z*_*i,j*_ = 1| *x*_*i,j*_, *ϕ*). The average of these scores, 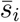, becomes the prediction for sample *i*. In CSNN-Reg, these scores serve as the output. For CSNN-Class, the scores are rescaled to determine a threshold for the classification.

**Fig. 2:**
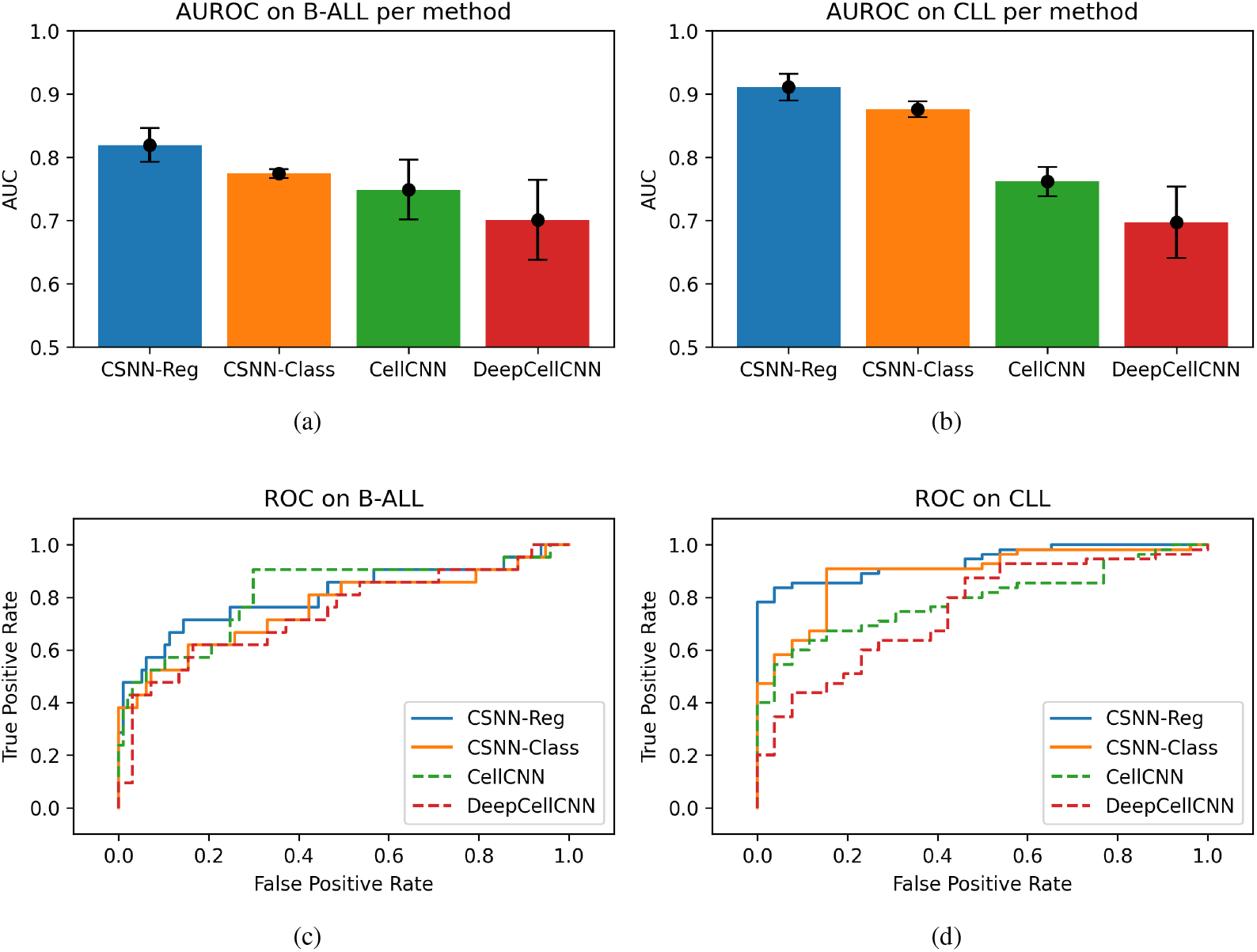
Model performance of sample-level leukemia classification. (a, b) The average of the testing scores for each model class on the B-ALL (a) and CLL (b) datasets, with the error bars representing the standard deviation between the N testing scores. (c,d) The ROC curve for the model with the highest scoring training AUC for each model class out of the N tests on the testing set of the B-ALL (c) and CLL (d) datasets.

In addition to performance on sample-level diagnosis, the CSNN-Reg method was also assessed for its ability to determine leukemic cell proportions on both the B-ALL and CLL datasets. Excellent correlation between the predicted proportions and the proportions reported by the diagnostic lab is observed for the CLL dataset (Fig 3). A similar correlation between predicted and reported leukemic cell proportions was observed for the B-ALL dataset. However, it should be noted that the set of samples that were randomly picked for testing did not contain any samples with proportions in the 0.2-0.8 range and therefore the performance of CSNN-Reg on the B-ALL dataset in this range is determined by interpolation. Ablation tests were conducted to evaluate whether the method performance is improved by the density difference initialization and the post initialization fine-tuning. Results of the ablation tests can be found in *Appendix 3*.

**Fig. 3:**
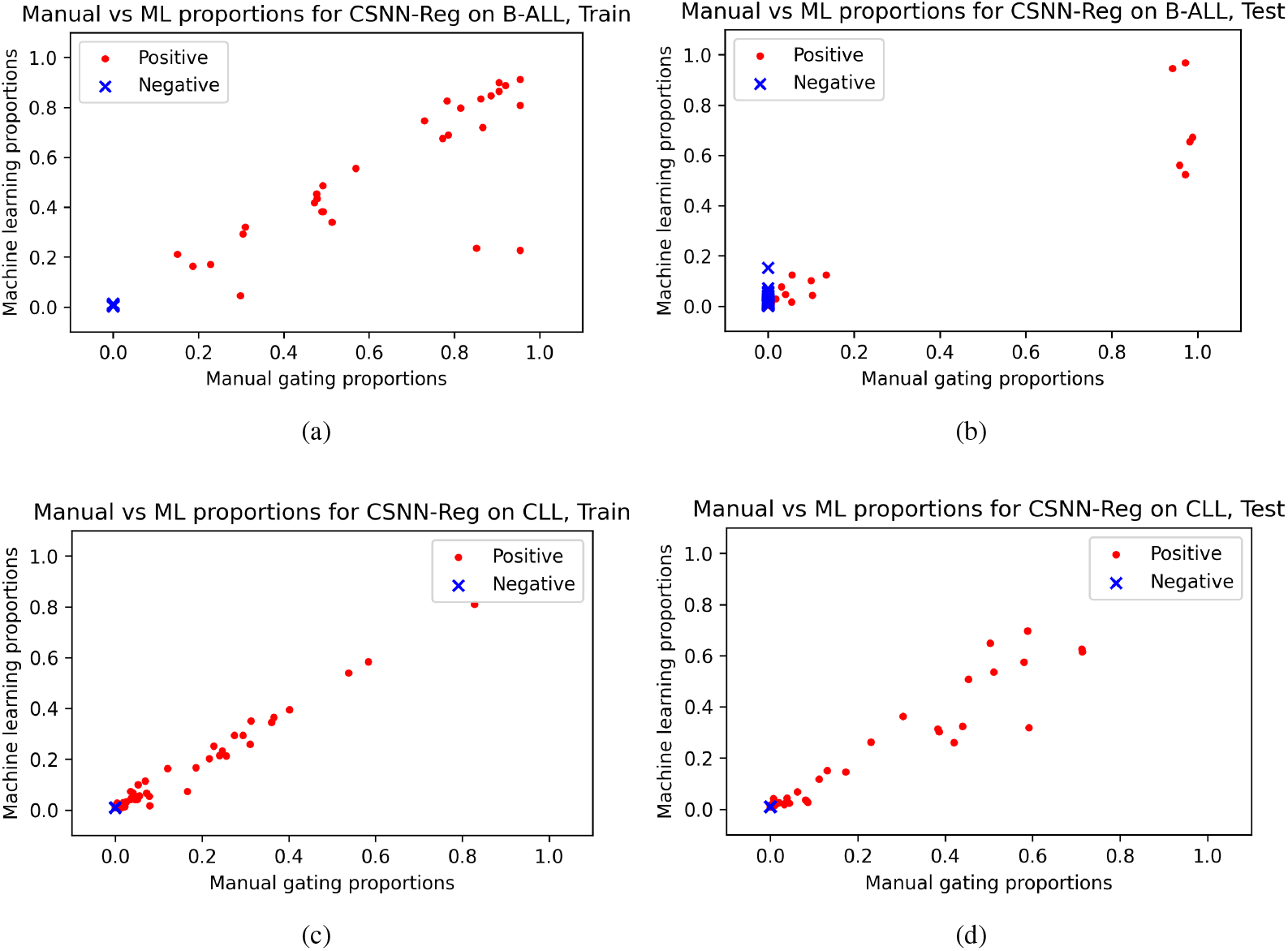
Comparison of machine learning- and manual gating-derived leukemic cell proportions. Using the best finetuned version of CSNN-Reg, the proportion of leukemic cells produced by the best finetuned version of SCNN-Reg and expert manual gating are compared the B-ALL (a, b) and CLL (c, d) training (a, c) and testing (b, d) data subsets.

#### 3.2.3 Biological interpretation of the identified cancer cells

The CSNN-identified pathologic cells are highlighted on the key 2D dot plots for visual examination and interpretation. We use the term “pathologic” to refer to cell populations that are related to the leukemic state, which can include both the leukemic cells themselves and any reactive “normal” cell population elicited by the presence of a leukemia in the patient that could be equally diagnostic and prognostic. Visual examination of the distributional shapes of antigen expression and locations of the identified pathologic cell populations in 2D dot plots was used to determine: a) if each CSNN-identified cell population has a natural shape on the 2D plot with a unimodal distribution of each marker, b) if the location of each CSNN-identified pathologic cell population matches a known leukemic cell phenotype, and c) if all leukemic cell populations seen on the 2D plots are successfully identified by the CSNN models.

Fig 4a shows the pathologic cells (in yellow) identified in the CLL dataset by the CSNN-Reg model from a few representative samples (results for all CLL positive samples can be found in *Supplementary Figure 2*). The CLL cell populations are highlighted across nine 2D dot plots that cover all the surface protein markers used in the reagent panel. This representative sample set consists of a negative case (row #1), positive cases with (row #2) and without (row #3) normal B cells, as well as CD38-negative (rows #2-3) and a mixture of CD38-negative and CD38-positive (row #4) CLL cases to illustrate within and between sample phenotypic heterogeneity. Many studies have previously reported the important role of CD38 in CLL prognosis [50, 42, 32, 35]. Fig. 4a clearly shows the capability and accuracy of the proposed CSNN model for identifying these important CLL phenotypic endotypes. Without being informed that the typical CLL phenotype is CD5^+^CD19^+^, the major pathologic cell populations identified by the CSNN-Reg were found to be CD5^+^CD19^+^. Without using clustering analysis to define cell populations up front, CSNN still successfully identified the cell populations with natural antigen expression distributional shapes and did not mix the normal CD19^+^ B cells with the CD5^+^CD19^+^ CLL cells (row #2), which can be difficult for traditional gating methods to cleanly separate. We also observed that the CSNN models do not require a prefiltering step needed by some existing methods to filter out debris/dead cells/doublets, such that any cell subsets in individual samples that differ between the CLL and non-CLL cohorts (e.g., the doublets in row #4, highlighted on FSC-A vs FSC-H) can be identified and used for classification.

**Fig. 4:**
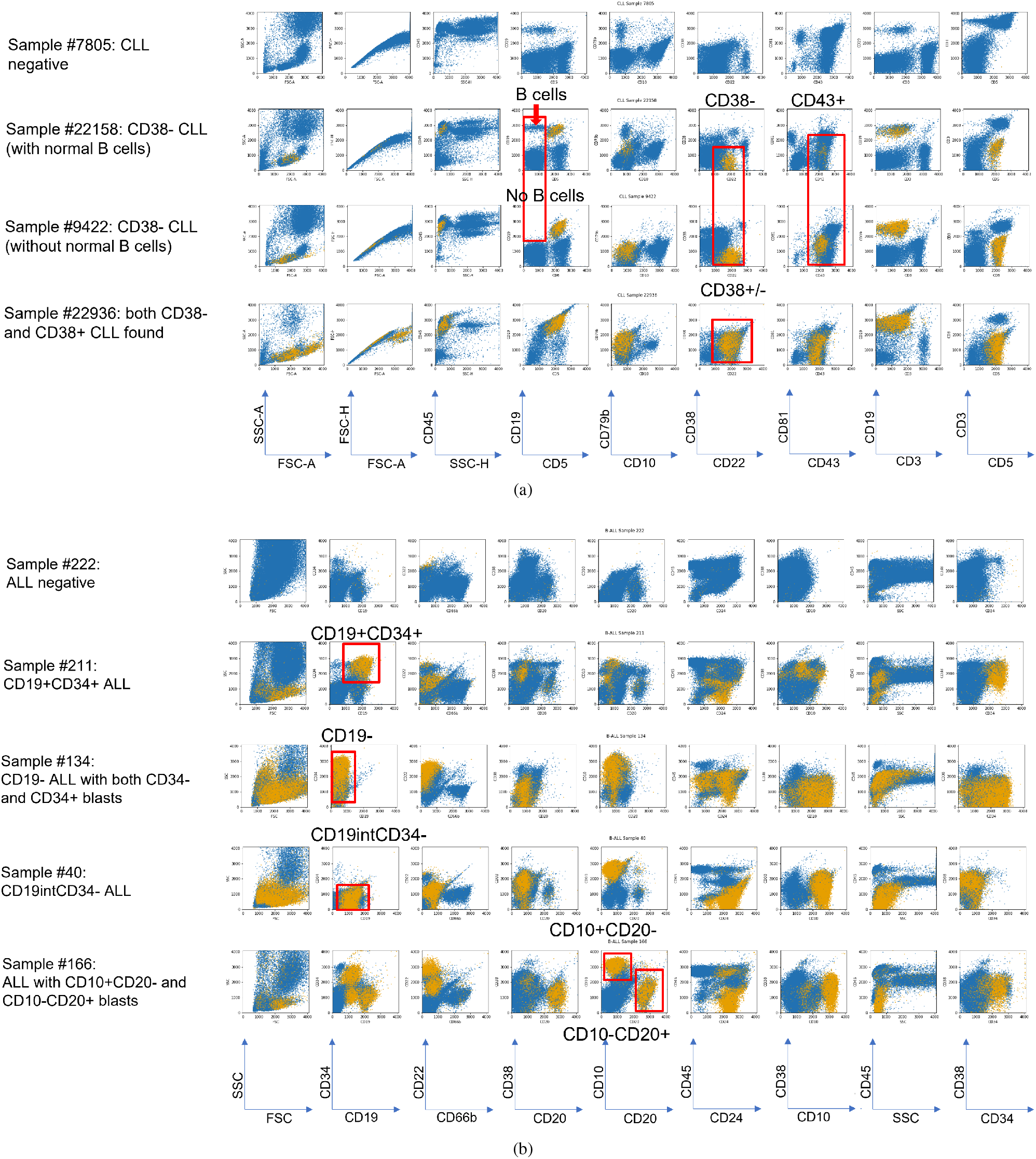
Visualization of the identified pathologic cells by CSNN-Reg. Pathologic cells identified in representative samples from the CLL (a) and B-ALL (b) datasets in nine 2D dot plots are colored yellow; non-pathologic cells are colored blue. Typical CLL cells are CD5^+^CD19^+^, while typical B-ALL cells are CD19^+^CD34^+^. Natural shapes of the identified pathologic cell clusters are produced by the CSNN model, without using clustering analysis. Phenotypic heterogeneity of the B-ALL cells can also be seen within and across the samples

The same visual assessment performed on the B-ALL dataset, highlighting the CSNN-identified pathologic cells from 5 representative samples (Fig 4b), including a negative sample (row #1), a typical CD19^+^CD34^+^ B-ALL sample (row #2), a CD19^−^ B-ALL sample (row #3, probably collected after CD19-targeted CAR-T therapy), an atypical CD19^*int*^CD34^−^ B-ALL case (row #4), and a B-ALL sample with at least 3 subtypes of blasts (row #5). Results from all B-ALL positive samples on CD19 vs CD34 can be found in *Supplementary Figure 3*. Expression of CD19, CD34, CD10, and CD20 in the B-ALL samples illustrates the phenotypic heterogeneity observed. These B-ALL phenotypic endotypes can be extremely challenging to identify using manual gating analysis due to this sample-to-sample heterogeneity. The CSNN model identified these phenotypically heterogeneous B-ALL cells at the single cell level by comparing all of the individual samples in the B-ALL positive and B-ALL negative cohorts, without requiring cell-level labels in the training data.

#### 3.2.4 Qualitative assessment and visual comparison of the results identified across competing methods

The results of the four different methods (CellCNN, DeepCellCNN, CSNN-Reg, and K-means) were visualized and compared for their identification of cancer cell phenotypes. The K-means clustering approach is an ad hoc independent, but fully interpretable, way of identifying cell clusters that can only be found in the cancer samples (method design can be found in *Appendix 4*). The complete set of 2D dot plots for visual comparisons of results of CSNN-Reg with the two baseline methods on all CLL and B-ALL samples can be found in *Appendix 5*. Fig. 5 shows the results of CellCNN (row #1), DeepCellCNN (row #2), CSNN-Reg (row #3), and K-means (row #4) on selected CLL (case #22936, Fig. 5a) and B-ALL (case #134, Fig. 5b) cases across nine different 2D dot plots (columns). The 2D plots of the baseline methods were generated using a probability cutoff = 0, because a cell should not be counted as a non-leukemic cell if it increases the likelihood of the sample being positive, and vice versa. Fig. 5 shows that both CellCNN and DeepCellCNN identified few leukemic cells using the probability cutoff = 0, indicating that the probability values output by the baseline methods cannot be directly used to predict whether a cell is leukemia-related or not, which is not too surprising, given that these two baseline methods were not designed for cell-level classification for blood cancer diagnosis.

**Fig. 5:**
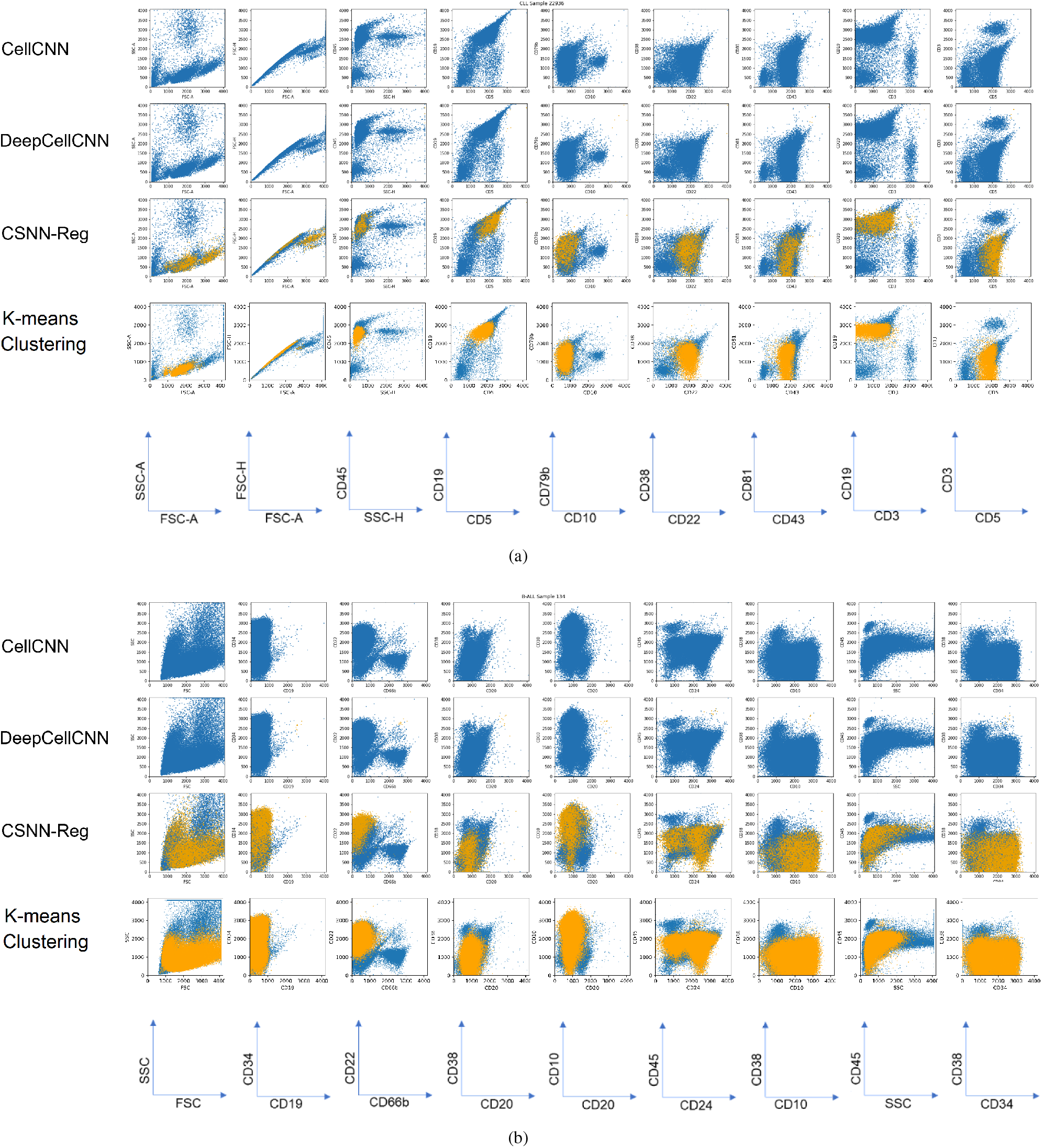
Visualization of pathologic cells identified by the different modeling methods. Leukemic cells identified from positive CLL case #22936 (a) and B-ALL case #134 (b) by CSNN-Reg (row #3) versus two baseline methods CellCNN (row #1) and DeepCellCNN (row #2) and an independent data clustering analysis using K-means (row #4) are highlighted in yellow with the rest of the cells in blue. Neither CellCNN nor DeepCellCNN could identify leukemic cells under their default settings.

#### 3.2.5 Interpretation of the neural network classification model using decision trees

In order to understand how CSNN was able to identify the heterogeneous leukemia-related cell subsets in individual samples, all B-ALL samples were pooled to construct a global decision tree to illustrate the classification paths of the cells in the pooled sample, based on the CSNN-output labels at the single cell level. Trees were then generated for each individual sample by calculating the cell-level statistics of the sample following the classification structure of the global tree, which preserved the tree layout for result comparison and interpretation across individual samples.

Using decision trees to interpret neural network analysis results is not new and has been discussed previously [11, 17]. However, Fig. 6 shows that a tree-based classifier can be adapted for not only interpreting the sample-level classification but also illustrating the phenotypic heterogeneity of B-ALL cells in individual patient samples. Three representative B-ALL positive samples were selected for visualizing the tree-based classification paths derived from the CSNN-identified cell-level labels side by side with the 2D dot plots that highlight these B-ALL cells (Fig. 6): case #134 (top) is a CD19^−^ B-ALL example, case #211 (middle) is a CD19^+^ B-ALL example, and case #166 (bottom) is an example that contains a mixture of CD19 negative and positive leukemia-related cells. Each node in the tree corresponds to a protein marker used in the reagent panel. The root of the tree is automatically selected during the tree construction process as the most informative feature for classification. In the pooled B-ALL sample, CD19 was identified as the root of the tree, which matches with our understanding of the patient cohort, in which some have been treated with the CD19-targeted CAR-T therapy and therefore the leukemic cells that have remained following therapy have lost expression of CD19. Our decision tree model, derived from the CSNN output at the single cell level, successfully identified the two major B-ALL subtypes in the patient cohort: naive CD19^+^ B-ALL and treatment-related CD19^−^ B-ALL.

**Fig. 6:**
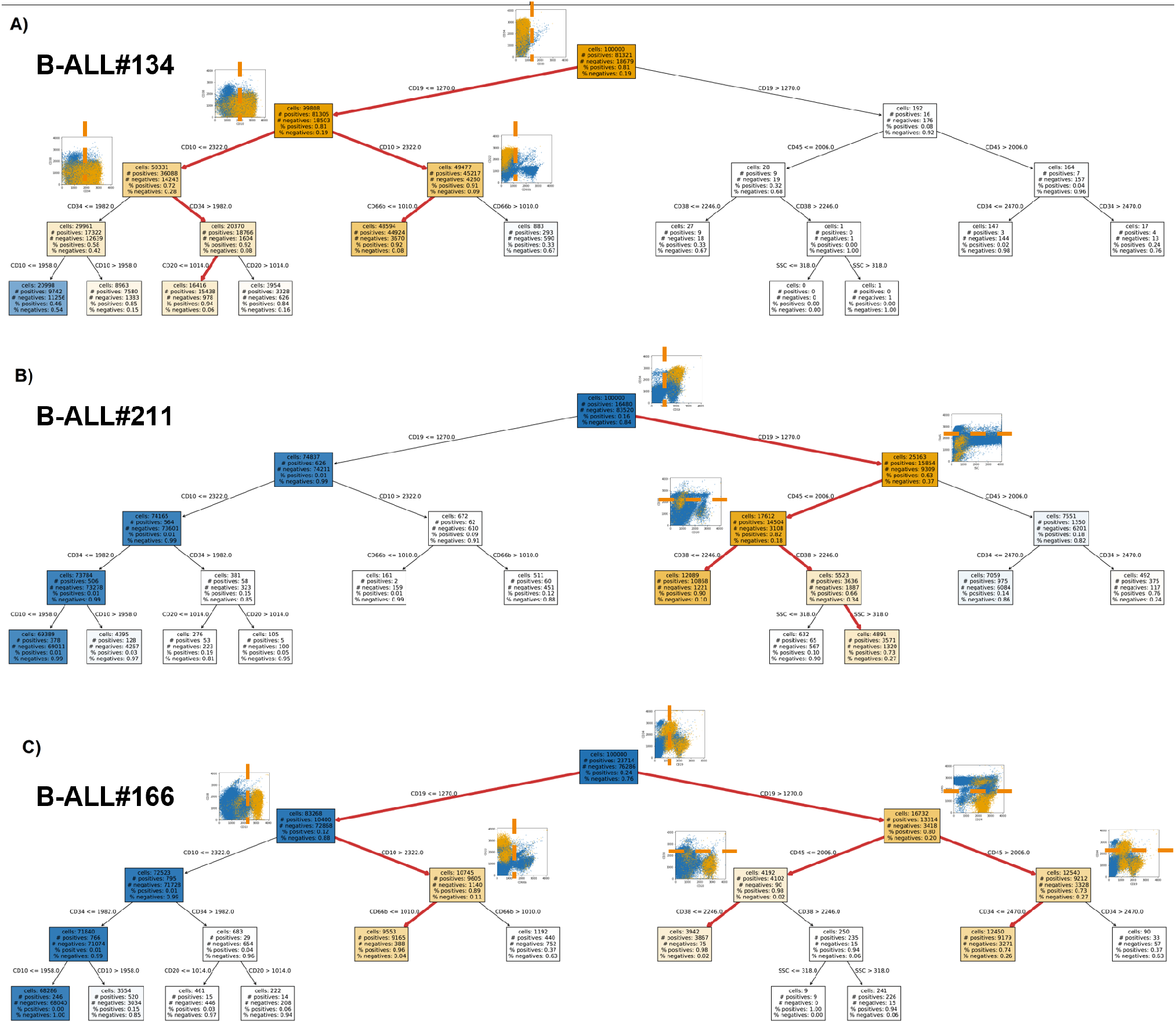
Neural network model interpretation using decision trees. 2D dot plots show the important tree nodes and highlight the leukemic cells identified and the expression cutoffs of the corresponding markers. Heterogeneity of B-ALL can be clearly seen in A) CD19^−^ B-ALL found in sample #134, B) CD19^+^ B-ALL found in sample #211, and C) both CD19^−^ and CD19^+^ B-ALL can be found in sample #166. The orange dotted lines in the 2D plots indicate the marker expression cutoffs identified by the decision tree classifier. Conceptually, each path highlighted in red can be thought of as corresponding to a traditional manual gating sequence with marker expression cutoffs determined in a data-driven manner.

Indeed, each path in the tree-based model leading to a B-ALL positive leaf node corresponds to a distinct B-ALL phenotype, potentially defining B-ALL endotypes (Fig. 6). In case #134, while all of the B-ALL cells are CD19 negative, the CD19^−^ B-ALL cell phenotypes can be further subdivided based on CD10, CD34, CD20, and CD66b expression. For case #121, while all of the B-ALL cells are CD19 positive, they can be further subdivided based on CD45 and CD38 expression, and SSC characteristics. The B-ALL cells in case #166 consist of three major subtypes: CD19^−^CD10^+^CD66b^−^, CD19^+^CD45^−^CD38^−^, and CD19^+^CD45^+^CD34^−^. Navigating along the decision tree provides an exploratory capability of identifying both known and novel leukemia-related cell phenotypes in individual samples, in a data-driven exhaustive way.

Finally, each tree path can also be interpreted as a manual gating strategy, with the marker expression cutoff values identified at the tree nodes defining the gating boundaries in the original marker space. In case #134, 1270 on CD19 is the cutoff for dividing the cells into CD19^+^ and CD19^−^. Visualization of the 2D dot plots confirms that the CD19 cutoff follows the natural boundary of the CD19 expression distribution. Similarly, the cutoffs of 2322 on CD10 and 1010 on CD66b for case #134 also follow the data distribution for separating positive from negative cells. In case #211, 2246 on CD38 derived from the CSNN output seems a perfect global gating cutoff for dividing the cells into CD38 negative and CD38 positive. Importantly, CSNN successfully calculated the local gate (the B-ALL cell population is highlighted in yellow), without relying on the global cutoff. The interpretation of these results is two-fold. First, the cell-level labels output by CSNN can be used to derive an accurate global cutoff for visual interpretation and validation. Second, the sample-level CSNN results provide for sample-specific cell classification that may deviate from the global cutoff identified in the pooled decision trees. This suggests that it could be very challenging to identify the B-ALL cell populations across all samples using traditional manual gating analysis. The same phenomenon was observed in case #166 on CD19 where the data and plots clearly show that there exists phenotypic heterogeneity of B-ALL within and between individual samples. While a global cutoff can be precisely identified based on the pooled data, the leukemic cells in individual samples needed to be identified using a “local gate” as predicted by CSNN. In clinical practice, the cutoffs and 2D dot plots output by CSNN along with the tree-based classification paths can be combined and converted into manual gating strategies for explainable validation by hematopathologists.

## 3. Additional findings

When examining 2D dot plots across individual samples, we noticed that CSNN was able to identify the leukemia-related cells with distinct atypical phenotype, which could be useful for cancer precision medicine. *Supplementary Figure 4* shows B-ALL sample #40 side by side with other five other B-ALL samples on plots of CD34 vs CD38, in which few typical CD34^+^ B-ALL cells are observed in Sample #40. However, the hematopathology report listed that the cancer burden of Sample #40 is 86.19%. *Supplementary Figure 5* compares sample #40 with the typical B-ALL case #211, where CSNN identified an atypical CD19^*int*^CD34^−^ leukemia-related cell population from sample #40 at 83.8%, matching the hematopathology review result. cellCNN and DeepCellCNN could not identify this cell subset in Sample #40.

Another finding from reviewing the 2D dot plots is that CSNN was able to capture cell populations with natural protein expression distributions, similar to what unsupervised clustering analysis can do. In contrast, manual gating analysis, decision-tree classification, and statistical biomarker identification methods often involve abrupt expression cutoffs that do not reflect natural expression gradients. It is important to note that the models produced by CSNN do not generate or rely on any geometric shape of gates but identify the leukemic cell populations as continua. The CSNN models can identify multiple fine-grained (hyper)regions that differ between the cancer and non-cancer cohorts, which allow the identification of complex classification patterns not easily captured through sequential gating methods.

A third finding is that the experiment results of CSNN show clear improvements in tagging cell-level labels, as it explicitly model whether each cell contributes to a sample-level classification as having leukemia or not. In contrast, other approaches such as DeepCellCNN [17] define the label of a cell as a product of its classification, by amplifying the cell to be 5% of a sample, followed by calculating the difference in the classification likelihood, resulting in a less robust heuristic, as cancer heterogeneity cannot be explained through amplifying the same single cell.

## 4 Model limitations and future extensions

A challenge that arises when evaluating the performance of these algorithms is the lack of ground truth annotations at the cell level. Although manual gating analysis can generate cell type labels of individual cells, they are only for known cell types and their precise accuracy is questionable due to the subjective manual operation. The only reliable evaluation metric is the classification error of the whole sample, based on the diagnostic labels of each sample. Therefore, to assess cell level classification we rely on visual examination of the cell populations on the original 2D plots in order to confirm that they match the known leukemic cell phenotypes. To improve this situation, we designed an independent data clustering approach (Appendix 4) to identify the cells that can only be found in the cancer samples. This allows us to compare and qualitatively confirm that the pathologic cells identified by CSNN are leukemia related. As CSNN is a probabilistic model, the model assigns a probability to each cell of being leukemia related. For non-leukemic cells, the probability values can be extremely low, usually around 1%; however, they are seldom equal to 0. The aggregated score is an estimator for the expected value of the tumor burden,

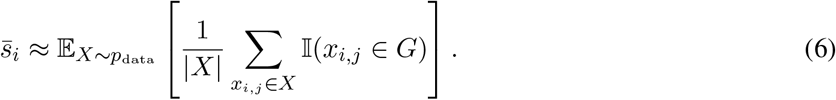

As the probability of a cell being a leukemic cell is usually non-zero, the above equation will return a non-zero value, even when the model has not found any cells that are likely to be related to the leukemia. In this case, a small number of cells may be classified as leukemic using a discrete threshold even for a non-cancer sample, as long as the diagnosis of the sample is correctly predicted by the model.

As the focus is on minimizing a global objective, the training algorithm might overlook small (<1%) populations of cells in favor of correctly classifying the majority of the samples. This issue prevents the model from identifying specific cell populations that are found in only a small number of samples. Similarly, the model will have difficulty classifying samples that have very small numbers of cancer cells, e.g., minimum residual diseases (MRD), especially if these MRD samples are not included during training. A localized training loss objective designed specifically for identifying MRD samples could help solve this problem. The proposed model could also benefit from identifying and modeling cell populations as a hierarchy. Then each cell population, instead of individual cells, could be scored. Most machine learning models are discriminative, without modeling the cell populations in a generative way, as such, can be hard to interpret. By separating each cell into a cluster and then classifying these clusters individually, both performance and interpretability of the model may improve.

An immediate next step is discrepancy analysis. For discrepant predictions for sample diagnosis and cancer burden, we will need to plot the identified leukemic cells on the original sequential gating paths for hematopathology review. For each false positive case, we plan to investigate whether the subject eventually develops leukemia at a later time point when further clinical data are available and approved for research use.

## 5 Conclusion

The most important feature of the CSNN model is its capability of simultaneously predicting the diagnosis of a sample and identifying pathologic cells, even with phenotypic heterogeneity. Existing machine learning methods for FCM data analysis either are not designed for blood cancer diagnosis or do not identify and validate the cancer cells from individual samples for result interpretation. In order to demonstrate this capability and assess the performance of CSNN, we designed a suite of interpretation and validation approaches for comparing the CSNN results to independent clustering analysis, known patient endotypes, diagnostic labels, and expert-identified cancer burden, in addition to hematopathology review of the CSNN-identified cancer cells on original 2D dot plots. Using two independent experiments on CLL and B-ALL, we showed the superiority of CSNN over the existing representative neural network modeling approaches for blood cancer diagnosis. The proposed neural network model is generally applicable to other types of discriminative single cell data analysis.

## Data Availability

The source code of the CSNN can be downloaded at GitHub: https://github.com/erobl/csnn. The de-identified B-ALL dataset is publicly accessible on FlowRepository under accession FR-FCM-Z6YK. The CLL dataset was converted to TXT format during de-identification, which can be downloaded at GitHub: https://github.com/JCVenterInstitute/DAFi-gating/tree/master/CSNN/CLL_TXT. 2D dot plots for comparing CSNN with other methods can also be found at GitHub: https://github.com/JCVenterInstitute/DAFi-gating/tree/master/CSNN/Comparison_with_CellCNN_DeepCellCNN.

## Data Availability

https://flowrepository.org/id/FR-FCM-Z6YK

https://github.com/JCVenterInstitute/DAFi-gating/tree/master/CSNN

## Acknowledgements

We thank Dr. Holden Maecker for connecting us with the Stanford Diagnostic Lab for performance assessment of the proposed models. This work has been partially supported by the FlowGate project (NCATS U01TR001801) funded by the National Center for Advancing Translational Sciences at the U.S. National Institutes of Health and an Investigator Sponsored Study Award (Protocol 68754391) from Becton, Dickinson and Company (BD).

## Notes

### Competing Interest Statement

Dr. Yu "Max" Qian has consulted for Moderna, Inc.

### Author Declarations

Datasets used in the study were collected using standard diagnostic protocols at University of California, San Diego and Stanford University. All necessary patient/participant consent has been obtained and the appropriate institutional forms have been archived. All datasets were de-identified before data transfer and analytics. Internal Review Board (IRB) of University of California, San Diego and Internal Review Board (IRB) of Stanford University have provided approval for the research work.

