## Appendix for "A cell-level discriminative neural network model for diagnosis of blood cancers"

### A Appendix

#### A.1 Specific hyperparameters

For our logistic method, the tuning hyperparameters used were the learning rates ($10^{-3}$, $10^{-4}$, $10^{-5}$, $10^{-6}$), the $w$ parameter (0.75, 0.85, 0.90, 0.95, 0.99, 0.995), and also an architecture search, where we test two kinds of architectures for the $s$ function, one with $\ell$ dense layers (1, 2, 3), and of size $n$ (3, 6, 10, 15). The other kind of architecture is a funnel that starts at size $2^n$ (for $n = 8, 4, 2$), half at each step and ends at size 1.

For our second proportion-based method, the hyperparameters tuned were the learning rate ($10^{-3}$, $10^{-4}$, $10^{-5}$, $10^{-6}$), negative penalty $\lambda$ (1, 5, 15, 30), and the same architecture search as the logistic method for $s$.

For DeepCellCNN, we use the following hyperparameters: the learning rate ($10^{-3}$, $10^{-4}$, $10^{-5}$, $10^{-6}$), and then varying the architecture, with an option for the hidden layers (3, 6, 10, 1), the embedding dimension size (3, 6, 10, 15), and the hidden layer of the head of the neural network (3, 6, 10, 15).

For CellCNN we use the following hyperparameters: the learning rate ($10^{-3}$, $10^{-4}$, $10^{-5}$, $10^{-6}$), then the architecture, with the number of filters (3, 6, 10 and 15), and the dropout (from 0.1 to 0.9 with a step every 0.1).

#### A.2 Interpretation of cell-level scores as gates

Traditional manual gating methods use a similar strategy to the method we just described: humans look at 2D plots of marker values and draw a box where they believe pathogenic cells are located for that pair of markers. This is repeated using additional 2D plots, filtering out the cells outside of the bounding box at each step. In the end, human gaters end up generating a similar function $s'$, where

$$s'(x; G) = \begin{cases} 1 & \text{if } x \in G \\ 0 & \text{otherwise} \end{cases}, \tag{1}$$

where $G$ is a set that describes the space after all the filtering done by a human is applied. We can interpret the score of a cell $s_{i,j} = s(x_{i,j}, \phi)$ as the probability of it being a pathogenic cell. This is because when using $L_{MSE}$ and $L_{LL}$ as our mappings when scoring a whole sample, the final probability is proportional to the mean of the probabilities in the sample. If we consider the set $G$ such that

$$G = \{x | s(x, \phi) > 0.5\}, \tag{2}$$

The function $s$ represents the human filtering done through $s'$. $G$ can also be interpreted as the location in marker space where pathogenic cells are likely to be found, since it is the region where $s$ has a high activation, and high activation is proportional to a high probability score. These scores can also be any number between 0 and 1, representing the probability that a cell is pathogenic. In practice, these are usually close enough to 0 or 1, to the point where they're all essentially non-pathogenic or pathogenic.

It is worth noting that $s$ has more representation power than $s'$, this is because while $s$ is represented by a cell scoring neural network, which allows it to have any arbitrary shape, $s'$ is represented by a hierarchy of 2D boxes, which can only be reduced to $n$ dimensional boxes. This means that, if we consider the case of two markers $A$ and $B$, where cancer cells are both $A^-B^-$ and $A^+B^+$ and healthy cells $A^+B^-$ and $A^-B^+$, a single 2D box cannot filter cancer cells based on these markers, whereas a neural network has no trouble making these distinctions.

#### A.3 Ablation study of the method

Three ablation tests were conducted to determine if the CSNN methods are improved by either the density difference initialization or the post initialization fine-tuning. The first test consisted of running the initialization score as a classification score without involving any neural network - denoted as Init-only. The other two ablation tests used CSNN-Reg and CSNN-Class without the density difference initialization - denoted as CSNN-Reg-noinit and CSNN-Class-noinit. The tests were run for 5 times and the best scores are reported in Table 1. These results indicate that both the initialization and the post-initialization fine-tuning have a significant impact on the accuracy of the models.

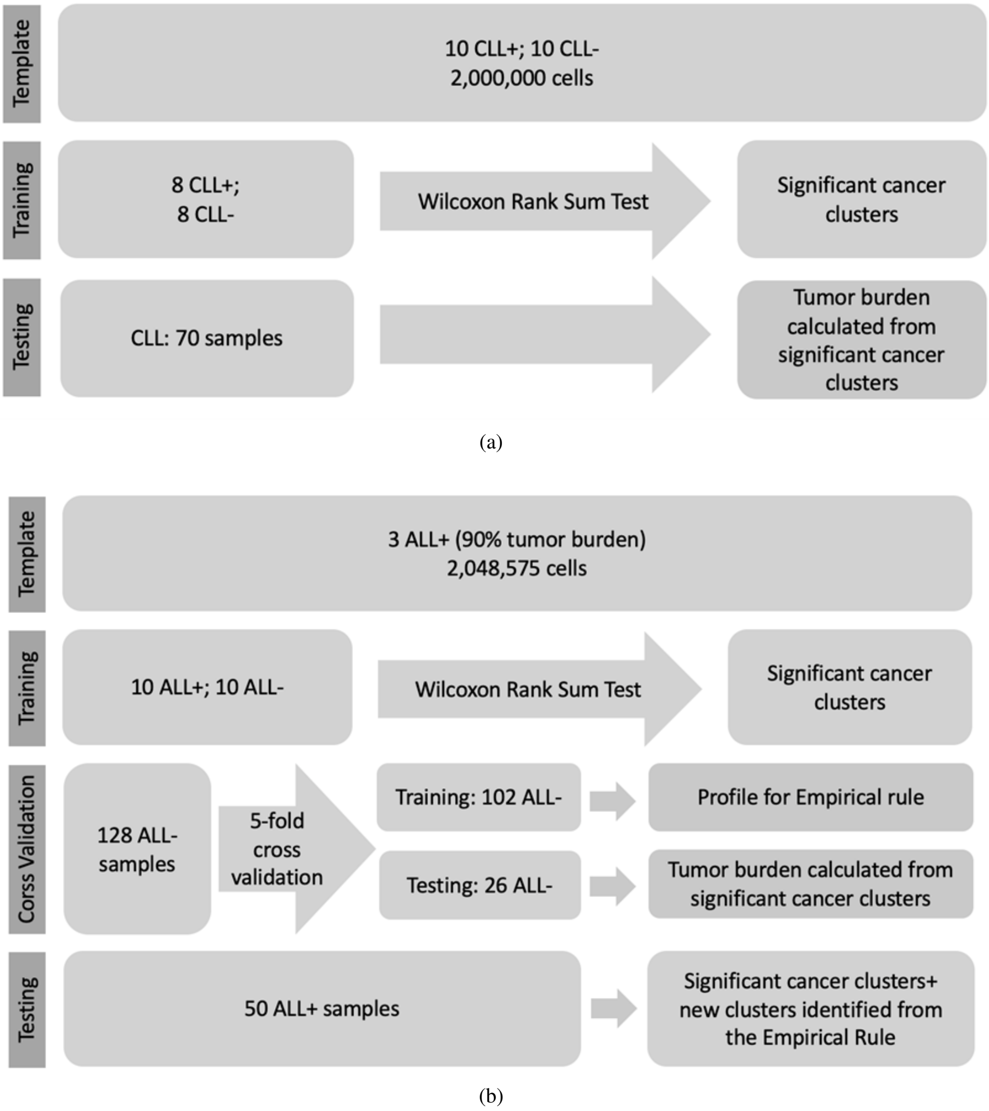


Fig. 1: Workflow of K-means clustering identification of cancer cells from (a) CLL and (b) B-ALL datasets.

#### A.4 Clustering-based independent analysis

We designed an ad hoc data clustering application to identify the different clusters of cells between the non-cancer and cancer samples (Fig. 1). A simple K-means clustering was applied to the original data space without any "black box" feature transformation or preprocessing steps. Due to the long runtime of K-means on large datasets, we selected and merged a small number of samples to create a template for identifying the optimal positions of the cluster centroids using 300 K-means iterations. Then we applied these centroids as initial seeds to generate final clustering results for the other samples, using only two K-means iterations. K=100 (i.e., 100 clusters) was used to overpartition the data so that both abundant and rare cell populations can be identified for being compared between the cancer and non-cancer cohorts. The clusters that were found in the cancer cohort only will be output as "pathologic cells". Specifically, we selected 10 CLL positive samples with around 50% tumor burden and

Table 1. Area under the ROC for each ablation and result.

| | B-ALL | CLL |
|---|---|---|
| CSNN-Reg-noinit | 78.1% | 91.2% |
| CSNN-Reg | 79.1% | 94.4% |
| CSNN-Class-noinit | 76.3% | 78.0% |
| CSNN-Class | 77.6% | 76.3% |
| Init-only | 72.8% | 79.3% |

10 CLL negative samples to create the CLL template. As the ALL files are much larger than the CLL, 3 ALL positive files with 90% or larger tumor burden were pooled to create the ALL template.

We used the nonparametric Wilcoxon Rank Sum test to select the clusters that are significantly amplified in the cancer cohort with a p-value cut off = 0.05. These clusters are the "global cancer clusters" identified at the cohort-level, i.e., they are seen in most of the cancer samples. However, it is known that the B-ALL has much larger phenotypical heterogeneity than CLL, which requires us to create an additional rule to identify the sample-specific pathologic cells. We defined an empirical rule based on our observation: if the proportion of a cluster in a testing sample is larger than the average proportion + 3 * standard deviation of the same cluster in the healthy samples of the training set, this cluster is regarded as a cancer cell cluster, no matter this cluster is found in other cancer samples or not. A 5-fold cross-validation was used to identify the average proportion of each cluster in the healthy cohort. The remaining 50 ALL positive samples were reserved as testing set before clusters in each of them were compared with the average profile built from the healthy cohort. Finally, both the global and the sample-specific cancer cell clusters were output as the predicted cancer burden. The stepwise process, including numbers of training, validation, and testing samples for CLL and B-ALL can be found in Fig 1. The predicted cancer burden was compared with the known cancer burdens from expert manual gating analysis, with Pearson's correlation coefficient calculated (Fig. 2).

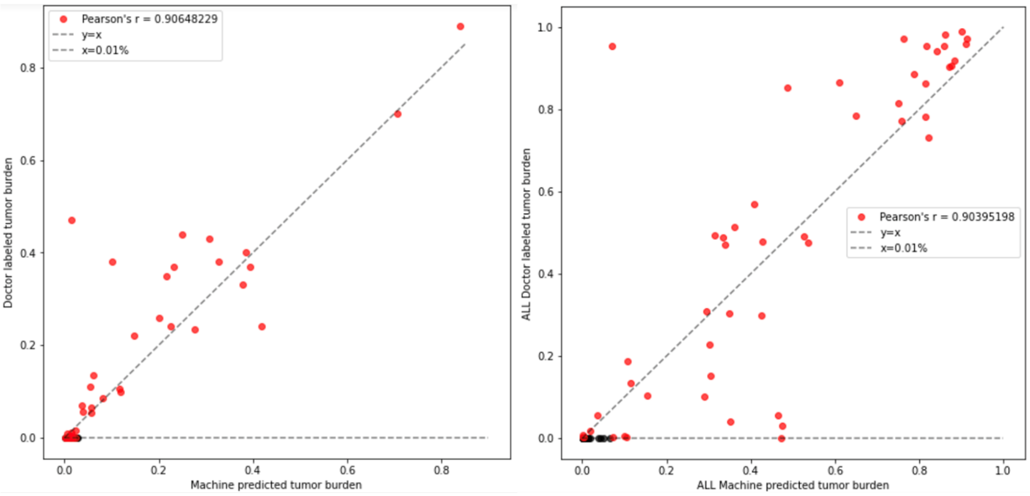


(a) (b)

Fig. 2: Pearson's correlation between proportions of cancer cells identified by K-means clustering (X-axis) and expert manual gating analysis (Y-axis) for samples in the testing set of (a) CLL and (b) B-ALL.

#### A.5 Comparison of results of CSNN with other approaches

2D dot plots for visually comparing CSNN with CellCNN and DeepCellCNN across all CLL and B-ALL samples can be found at GitHub:

https://github.com/JCVenterInstitute/DAFi-gating/tree/master/CSNN/Comparison_with_CellCNN_DeepCellCNN.
